# Patterns of Post-Traumatic Stress Disorder and Associated Cognitive Factors Among Flood Victims in Hanang District, Tanzania

**DOI:** 10.64898/2026.09.01.26361894

**Authors:** Elias Manaseh Mwana, Leonard Katalambula, Basiliana Emidi, Azan Nyundo

**Affiliations:** University of Dodoma, Department of Public Health, Dodoma, Tanzania; National Institute for Medical Research, Dodoma Medical Research Centre, Dodoma, Tanzania

**Keywords:** PTSD, floods, disaster mental health, cognitive factors, trauma, Tanzania, Hanang district

## Abstract

**Background:** Floods are among the most devastating natural disasters worldwide and are increasingly associated with adverse mental health outcomes, particularly Post-Traumatic Stress Disorder (PTSD). In December 2023, Hanang District in northern Tanzania experienced catastrophic mud floods that resulted in extensive loss of life, destruction of property, displacement of households, and disruption of livelihoods. While emergency humanitarian responses focused primarily on physical needs, limited evidence exists regarding the long-term psychological consequences among survivors. Therefore, this study aimed to determine the patterns of PTSD manifestations and assess cognitive factors associated with PTSD symptoms among flood victims in Hanang District, Tanzania.

**Methods:** A community-based cross-sectional study was conducted among 360 flood victims one year after the disaster. PTSD symptoms were assessed using the PTSD Checklist for DSM-5 (PCL-5). Descriptive statistics summarized PTSD severity, while chi-square tests and regression analyses examined associations between socio-demographic characteristics and PTSD manifestations. Cognitive factors were assessed based on participants’ exposure to traumatic experiences and perceptions of traumatic events.

**Results:** The mean PCL-5 score was 39.2 (SD = 20.6), indicating a high burden of psychological distress. Approximately 45% of respondents had severe PTSD symptoms (PCL-5 ≥45), while another substantial proportion demonstrated moderate symptom severity. PTSD manifestations varied significantly by geographical location (p < 0.001), household income (p = 0.011), and marital status (p = 0.002). Age positively predicted PTSD severity (β = 0.019, p = 0.001), whereas household income negatively predicted symptom severity (β = −0.297, p = 0.001). Exposure to natural disasters constituted the predominant cognitive factor, with 45% directly experiencing the flood and 38.3% witnessing the event. Exposure to secondary traumatic experiences through witnessing or learning about violent events was also common. Cognitive trauma exposure demonstrated a significant association with PTSD symptoms (χ², p < 0.001).

**Conclusion:** PTSD remains highly prevalent among flood survivors in Hanang district. Both direct and indirect trauma exposure significantly contributed to PTSD manifestations. Comprehensive disaster recovery programmes should integrate trauma-focused psychological services, cognitive behavioural interventions, routine PTSD screening, and community-based psychosocial support alongside socioeconomic recovery initiatives.

## Introduction

Natural disasters are among the leading causes of mortality, displacement, and psychological morbidity worldwide. Their frequency and intensity have increased over recent decades due to climate change, rapid urbanization, environmental degradation, and population growth in disaster-prone areas (Intergovernmental Panel on Climate Change [IPCC], 2022; United Nations Office for Disaster Risk Reduction [UNDRR], 2023). Floods account for nearly half of all weather-related disasters globally and disproportionately affect low– and middle-income countries (LMICs), where limited infrastructure, poverty, and constrained healthcare systems increase community vulnerability (Centre for Research on the Epidemiology of Disasters [CRED], 2024). Although the immediate consequences of floods include deaths, injuries, destruction of infrastructure, and economic losses, the long-term psychological impacts often persist long after the physical damage has been repaired. One of the most common mental health disorders following disasters is Post-Traumatic Stress Disorder (PTSD). According to the *Diagnostic and Statistical Manual of Mental Disorders, Fifth Edition (DSM-5)*, PTSD develops following exposure to actual or threatened death, serious injury, or sexual violence, either through direct experience, witnessing the event, learning that it occurred to a close family member or friend, or repeated exposure to traumatic details (APA, 2022). PTSD is characterized by four major symptom clusters: intrusive recollections of the traumatic event, persistent avoidance of trauma-related stimuli, negative alterations in cognition and mood, and marked alterations in arousal and reactivity. If left untreated, PTSD can lead to chronic psychological impairment, depression, anxiety disorders, substance misuse, poor physical health, reduced quality of life, and impaired social and occupational functioning (Bryant, 2019; Koenen et al., 2017).

Flood disasters are particularly associated with elevated PTSD prevalence because survivors are frequently exposed to multiple traumatic experiences simultaneously. These include witnessing deaths, sustaining personal injuries, losing family members, destruction of homes and livelihoods, displacement, food insecurity, and prolonged uncertainty during recovery (Liu et al., 2021; Tang et al., 2017). A systematic review by Goldmann and Galea (2014) reported that PTSD prevalence following natural disasters ranges widely depending on disaster severity, population characteristics, timing of assessment, and access to psychosocial support, with estimates ranging from less than 10% to over 50% among directly affected populations. Similarly, studies conducted after floods in China, Pakistan, India, Australia, and the United States consistently report that PTSD remains prevalent months or even years after the disaster, particularly among individuals experiencing repeated trauma exposure and inadequate social support (Neria et al., 2008; North & Pfefferbaum, 2013).

In developing countries, the psychological consequences of floods are often exacerbated by limited access to mental health services, shortages of trained mental health professionals, weak emergency response systems, and widespread socioeconomic inequalities (WHO, 2022). Survivors frequently face prolonged displacement, unemployment, food insecurity, and disruption of community support systems, all of which increase vulnerability to persistent psychological distress (IASC, 2021). Furthermore, stigma surrounding mental illness and low mental health literacy often discourage affected individuals from seeking professional care, resulting in untreated PTSD and prolonged suffering (WHO, 2022). These challenges are particularly pronounced in many sub-Saharan African countries, where mental health services remain under-resourced despite increasing exposure to climate-related disasters. Across Africa, climate change has substantially increased the occurrence and severity of floods, droughts, and other extreme weather events (IPCC, 2022). Flood disasters in countries such as Mozambique, Nigeria, Kenya, South Sudan, and South Africa have resulted in significant psychological consequences among affected communities, including PTSD, depression, anxiety, and complicated grief (WHO, 2023). However, disaster mental health research remains limited across much of the continent, with relatively few studies examining long-term PTSD outcomes or the cognitive mechanisms underlying trauma responses. Existing evidence suggests that poverty, repeated disaster exposure, displacement, limited access to healthcare, and inadequate psychosocial support are major contributors to poor mental health outcomes among African disaster survivors.

Tanzania has experienced increasing frequencies of floods and landslides over the past decade, affecting numerous regions including Dar es Salaam, Kilimanjaro, Morogoro, Kigoma, and Manyara. These disasters have caused extensive destruction of infrastructure, agricultural production, housing, and livelihoods while exposing affected communities to considerable psychological trauma. Nevertheless, disaster response efforts have traditionally focused on emergency rescue, shelter, food assistance, and disease prevention, with comparatively limited attention given to mental health recovery. Consequently, there is limited empirical evidence describing the prevalence, manifestations, and determinants of PTSD among disaster survivors in Tanzania. On 3^rd^ December 2023, Hanang District in Manyara Region experienced one of the most devastating flood and mudslide disasters in Tanzania’s recent history following intense rainfall originating from Mount Hanang. The disaster caused substantial loss of life, injuries, destruction of homes, schools, health facilities, roads, bridges, farmland, and livestock, while thousands of residents were displaced. The floods disrupted livelihoods, separated families, and created prolonged socioeconomic hardship for affected households. Although humanitarian agencies and the Government of Tanzania mobilized emergency relief efforts to address immediate physical needs, relatively little attention has been directed toward understanding the long-term psychological consequences among survivors.

Previous research has demonstrated that PTSD following disasters is influenced not only by the degree of disaster exposure but also by cognitive factors that shape how individuals interpret and process traumatic experiences. According to cognitive theories of PTSD proposed by Ehlers and Clark (2000), persistent PTSD develops when individuals process traumatic experiences in ways that generate an ongoing sense of current threat. Maladaptive appraisals, persistent negative beliefs about oneself and the world, intrusive memories, rumination, catastrophic thinking, and dysfunctional coping strategies contribute to the maintenance of PTSD symptoms. Individuals who directly experience disasters, witness traumatic events, or repeatedly encounter reminders of trauma are therefore more likely to develop persistent PTSD if maladaptive cognitive processing occurs. In addition to cognitive factors, demographic and socioeconomic characteristics including age, sex, educational attainment, marital status, household income, and previous exposure to traumatic events, have consistently been associated with PTSD severity following natural disasters (Brewin et al., 2000; Lowe et al., 2019). However, the relative contribution of these factors varies considerably across different cultural and geographical settings. Consequently, understanding locally relevant predictors of PTSD is essential for developing context-specific interventions that effectively address survivors’ psychological needs.

Most available studies have focused primarily on immediate humanitarian outcomes, while long-term mental health consequences remain poorly understood. Furthermore, little is known about how cognitive trauma exposure—including direct exposure, witnessing traumatic events, and learning about trauma experienced by others, influences PTSD among survivors of the Hanang floods. Addressing this knowledge gap is critical for informing disaster preparedness strategies, integrating mental health services into emergency response systems, and developing evidence-based psychosocial interventions that promote resilience and recovery among disaster-affected populations. Therefore, this study aimed to explore the patterns of Post-Traumatic Stress Disorder (PTSD) manifestations and assess the associated cognitive factors contributing to PTSD symptoms among flood victims in Hanang District, Tanzania.

## Methods Study

### Design

A community-based convergent parallel mixed-methods study was conducted approximately one year after the Hanang landslide-flood disaster. The quantitative component employed an analytical cross-sectional design to assess the patterns and severity of PTSD manifestations and the cognitive factors associated with PTSD symptoms. The qualitative component used in-depth interviews to explore survivors’ experiences, interpretations of the disaster, intrusive memories, perceived threat, helplessness, negative appraisals, coping responses, and other cognitive processes related to PTSD. Quantitative and qualitative data were collected during the same study period and analysed separately before being integrated during interpretation. This approach enabled statistical assessment of PTSD patterns and associated cognitive factors while also providing contextual understanding of survivors’ lived experiences.

### Study Area

The study was undertaken in Hanang District, Manyara Region, Tanzania, among communities directly affected by the December 2023 landslide and flood disaster. Hanang is predominantly rural, with agriculture and pastoralism being major sources of livelihood. The district had a population of 367,391 according to the 2022 Population and Housing Census. The December 2023 disaster resulted in 85 deaths, displacement of more than 5,600 people, and substantial destruction of homes, infrastructure and livelihoods. The study was conducted in Gendabi, Jorodom and Waretta wards, which represented communities with different levels of disaster exposure and post-disaster living conditions.

### Study Population

The study population comprised adult flood survivors aged 18 years and above who were residents of Hanang District during the floods disaster and were directly exposed to or affected by the landslide-flood event. Eligible participants were required to have experienced direct disaster exposure, such as displacement, destruction of their homes or property, injury, loss of livelihood, or witnessing death or serious injury, and to provide informed consent. Individuals who were not directly exposed to the disaster, had severe cognitive impairment that prevented meaningful participation, or declined consent were excluded.

### Sample Size and Sampling

A total of 360 respondents participated in the quantitative component. The sample size was determined using Cochran’s formula, assuming a 50% prevalence, 95% confidence level and 5% margin of error. After applying finite population correction to the estimated 5,600 disaster-affected individuals, the required sample was approximately 359 and was rounded to 360 participants. Participants were selected using stratified simple random sampling, with Gendabi, Jorodom and Waretta serving as the sampling strata. Proportional allocation resulted in 144 participants from Gendabi, 126 from Jorodom and 90 from Waretta. For the qualitative component, 20 flood survivors were purposively selected from the three wards to provide variation in PTSD symptom severity, disaster exposure, sex, age, social role and post-disaster circumstances.

### Data Collection

Data were collected during December 2024 and January 2025, approximately one year after the disaster. Trained research assistants conducted face-to-face interviews using structured questionnaires and semi-structured interview guides. Study instruments were translated from English into Kiswahili using forward and backward translation and were pre-tested before the main data collection.

The quantitative questionnaire comprised several components designed to comprehensively assess the demographic characteristics, traumatic experiences, and PTSD symptoms among flood victims in Hanang District. The questionnaire collected socio-demographic characteristics of respondents, including age, gender, marital status, education, occupation, household income, and number of dependents. It also assessed disaster exposure and previous traumatic experiences, including direct, witnessed, and indirectly experienced traumatic events. The Life Events Checklist for DSM-5 (LEC-5) was used to assess exposure to potentially traumatic events, while the PTSD Checklist for DSM-5 (PCL-5) was administered to assess the presence and severity of PTSD symptoms. In addition, the questionnaire included questions assessing cognitive responses to the traumatic event, particularly intrusive memories, negative appraisals, perceived threat, feelings of helplessness, and loss of control. Together, these components enabled the study to examine the relationship between demographic characteristics, traumatic exposure, cognitive responses, and the manifestation and severity of PTSD among flood victims in Hanang District. The PCL-5 is a 20-item measure of PTSD symptoms corresponding to DSM-5 criteria. Participants rated how much they had been bothered by each symptom during the previous month on a five-point scale ranging from 0 (*not at all*) to 4 (*extremely*), producing a total score ranging from 0 to 80. The instrument assessed four PTSD symptom clusters: intrusion, avoidance, negative alterations in cognition and mood, and alterations in arousal and reactivity.

For the qualitative component, 20 participants participated in in-depth interviews lasting approximately 45–60 minutes. Interviews explored survivors’ experiences of the disaster, intrusive memories, perceptions of threat and safety, negative interpretations of the event, feelings of helplessness or guilt, avoidance, coping strategies and social support. Interviews were conducted in Kiswahili, audio-recorded with consent, transcribed verbatim and translated into English for analysis.

### Data Analysis

Quantitative data were analysed using IBM SPSS Statistics version 26. Data were checked for completeness, consistency and missing values before analysis. Descriptive statistics, including frequencies, percentages, means, standard deviations, medians and interquartile ranges, were used to summarize participant characteristics and PTSD symptom scores. The patterns of PTSD manifestations were assessed by examining the distribution and severity of the four PCL-5 symptom clusters. Chi-square tests were used to assess associations between categorical variables and PTSD categories, while appropriate comparative tests were used for continuous PTSD scores. To assess cognitive factors associated with PTSD symptoms, multiple linear regression was used when the PCL-5 total score was treated as a continuous outcome. Where PTSD was categorized as probable PTSD versus no probable PTSD, multivariable logistic regression was used to estimate adjusted odds ratios and 95% confidence intervals. Potential confounders, including age, sex, marital status, education, socioeconomic characteristics and severity of disaster exposure, were considered in the multivariable models. Qualitative data were analysed thematically following Braun and Clarke’s six-phase approach. Coding focused on cognitive processes such as intrusive memories, maladaptive appraisals, perceived threat, helplessness, negative beliefs and avoidance. Quantitative and qualitative findings were subsequently compared and integrated to identify convergence, complementarity and divergence between statistical patterns and survivors’ lived experiences. Statistical significance was set at p < 0.05, with 95% confidence intervals reported where appropriate.

### Reliability and Validity

The reliability of the quantitative instrument was supported through the use of the validated PCL-5, standardized administration procedures, training of research assistants, translation and back-translation, and pre-testing. Internal consistency of the PCL-5 was assessed using Cronbach’s alpha. Validity was enhanced through expert review of the study instruments, use of the PCL-5 aligned with DSM-5 PTSD criteria, appropriate sampling procedures, and adjustment for potential confounding factors during multivariable analysis. For the qualitative component, credibility was enhanced through in-depth interviews, triangulation of quantitative and qualitative findings, field notes and review of emerging themes. Dependability was supported by maintaining an audit trail of data collection and analysis procedures, while confirmability was enhanced through reflexive documentation and use of participants’ accounts to support interpretations.

### Ethical Considerations

Ethical approval for the study was obtained from the University of Dodoma Research Ethics Review Board before commencement of data collection. The study was conducted in accordance with relevant national and international ethical principles governing research involving human participants, including the principles of respect for persons, beneficence, non-maleficence, and justice (World Medical Association, 2013). Permission to conduct the study was also obtained from the relevant district authorities and community leaders in Hanang District. Before participation, each respondent was provided with clear information about the purpose of the study, the procedures involved, potential risks and benefits, and their rights as participants. Written informed consent was obtained from all participants prior to data collection. Participation was entirely voluntary, and participants were informed that they had the right to decline participation or withdraw from the study at any time without penalty or loss of any services to which they were otherwise entitled. Given the sensitive nature of the study and the possibility that recalling traumatic experiences could cause emotional distress, interviews were conducted in private and respectful settings. Participants were allowed to pause or terminate the interview whenever they felt uncomfortable or distressed, and those who demonstrated significant psychological distress or required additional support were referred to appropriate psychosocial or mental health services. Confidentiality and privacy were maintained throughout the study by assigning unique identification codes to participants instead of recording their names on the research instruments. Electronic data were password-protected and access was restricted to authorized members of the research team. Audio recordings, where applicable, and interview transcripts were securely stored and handled only by authorized researchers. These measures were implemented to minimize potential psychological and social risks and to protect the privacy, dignity, and well-being of all participants throughout the research process.

## Results

### Characteristics of Study Participants

**Table 1** presents the demographic characteristics of the 360 respondents included in the study. More than half of the respondents were male (57.2%), while females accounted for 42.8%. The majority were married (65.3%), followed by single respondents (19.2%), widowed respondents (8.3%), and those who were divorced or separated (7.2%). Educational attainment was generally low, with 71.7% having completed primary education, 19.4% secondary education, 6.7% having no formal education, and only 2.2% having attained higher education. Farming was the predominant occupation (68.3%), followed by small business activities (14.2%) and casual labour (4.4%). Government employees accounted for 1.1%, while 9.2% were unemployed. Regarding household income, 47.2% of respondents reported earning less than TZS 50,000 per month, indicating substantial socioeconomic vulnerability. The majority of respondents (57.8%) had between 0 and 4 dependents, while 34.2% had 5–9 dependents, 6.9% had 10–14 dependents, and 1.1% had more than 14 dependents.

**Table 1.**
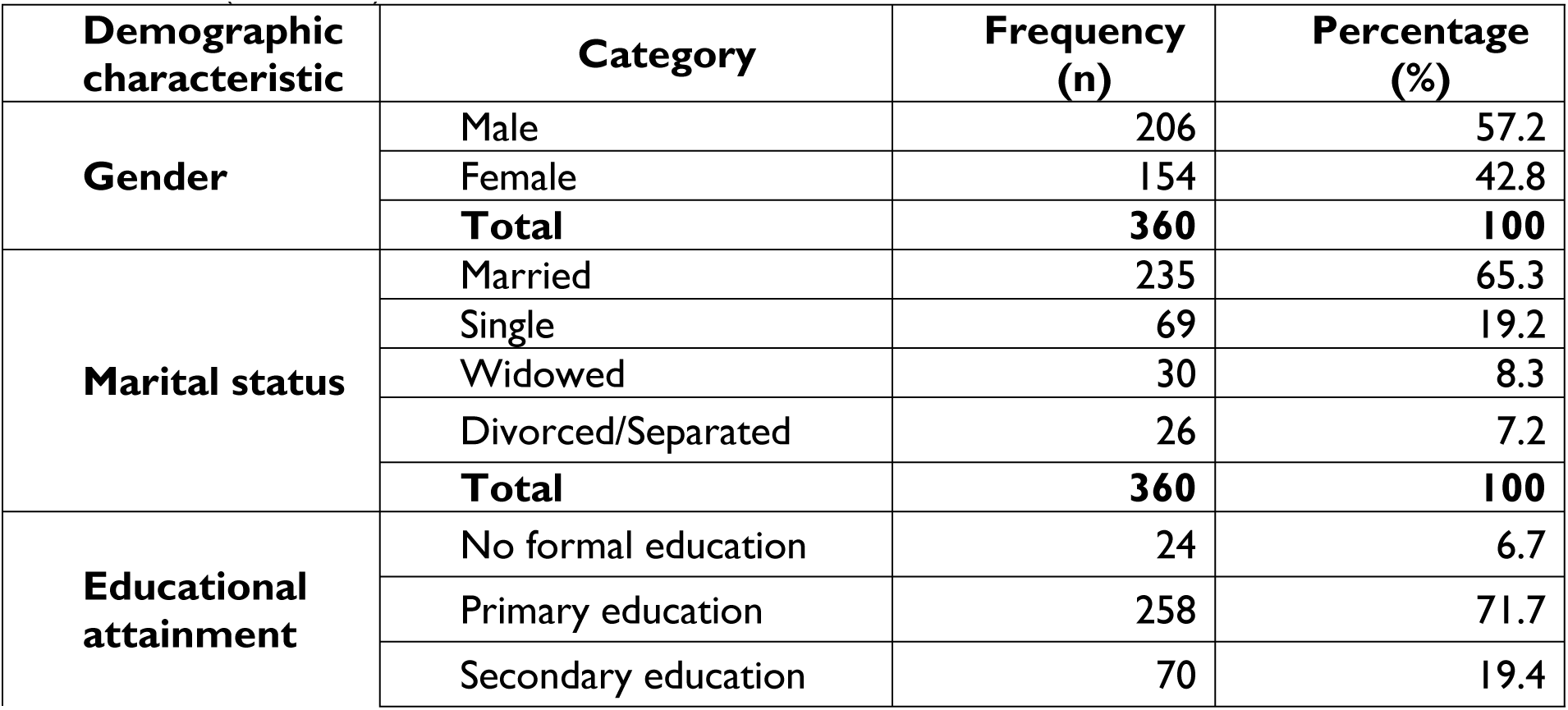
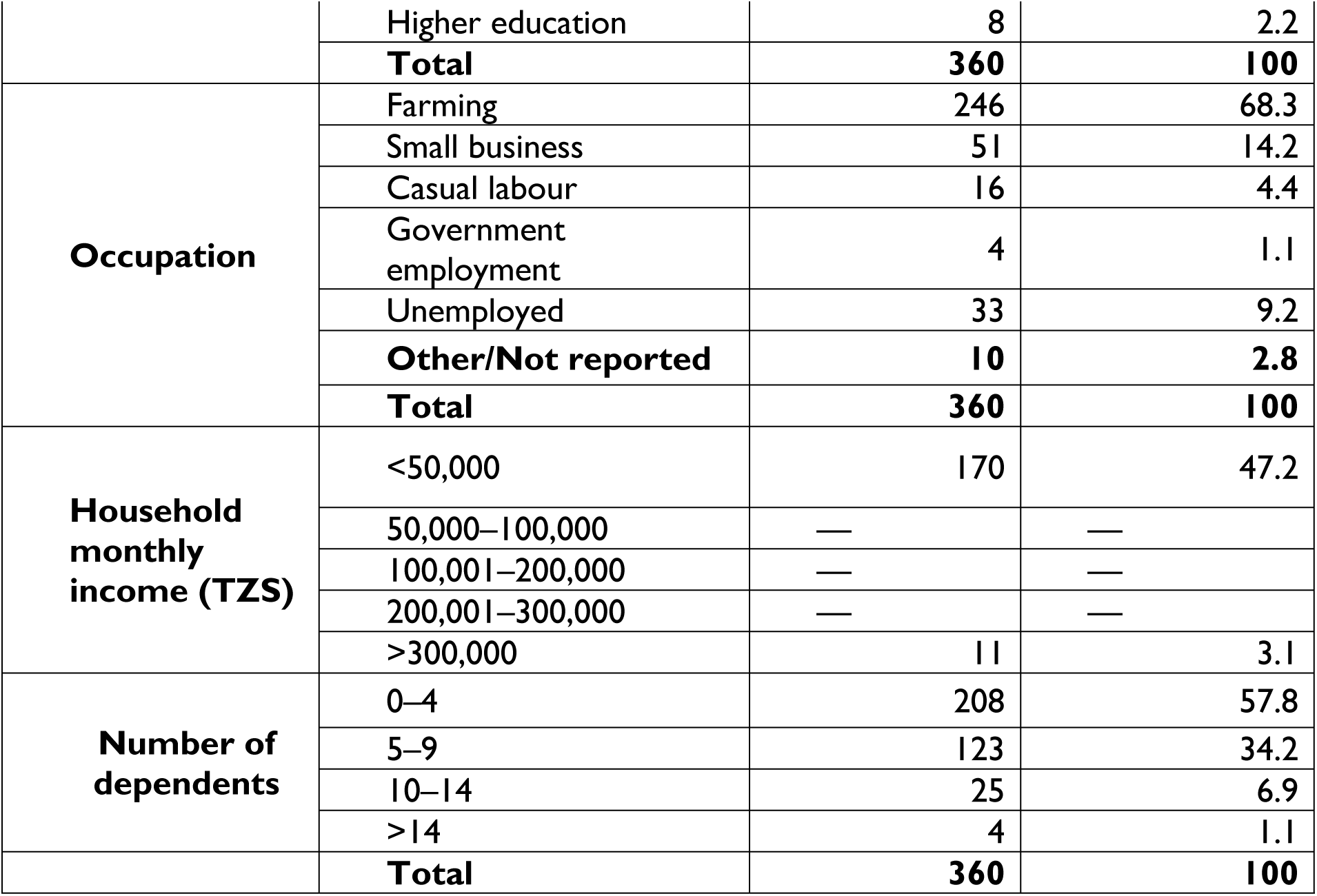
Demographic characteristics of respondents among flood victims in Hanang District, Tanzania (N = 360)

### Age Distribution

Regarding age of the respondents, the median age of the respondents was 39 whereas the average age was 41 years with a wide distribution of 18-78 years of the ages as shown in Table 2. This implies that the surveyed population is mostly adult though it includes both the younger and elderly ones. The rural economy and families mostly depend on middle-aged adults, and hence they are most prone to psychological pressure after disasters. According to a study by Qadir et al. (2024), the prevalence of PTSD was highest in adulthood in the age range between 31 and 45 because at this stage, people are responsible both as the breadwinners and caregivers. The occurrence of different age groups in the data also points out to needing age-specific trauma recovery programs in the district.

**Table 2:**
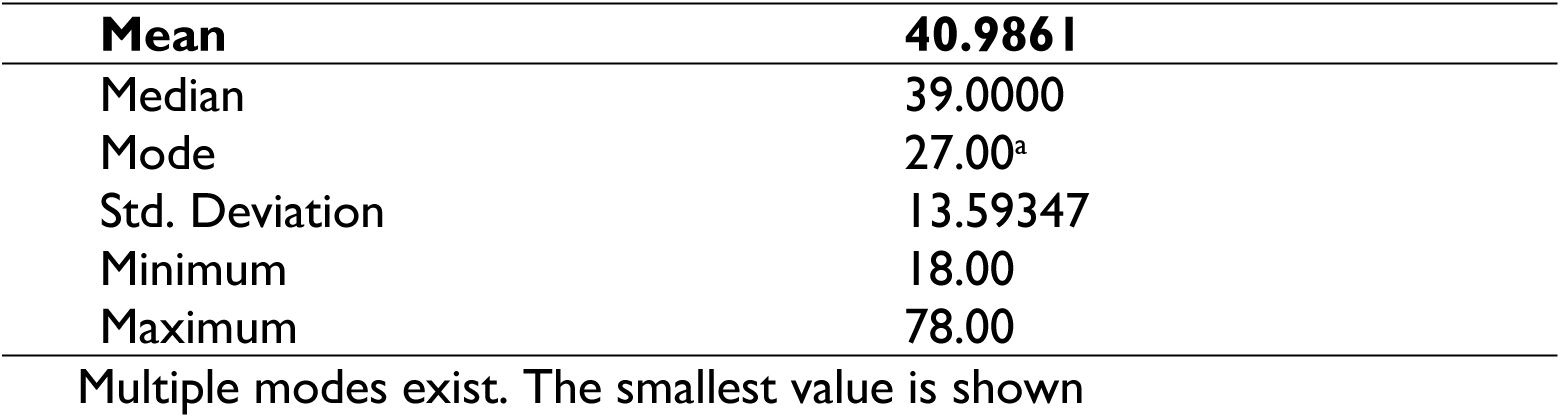
Age of respondents.

### Patterns of PTSD Manifestations among Flood Victims in Hanang District

PTSD severity was assessed using the Post-Traumatic Stress Disorder Checklist for DSM-5 (PCL-5) and categorized into five score groups: 0–14, 15–29, 30–44, 45–59, and >60. The distribution of PTSD severity was examined according to selected demographic and socioeconomic characteristics, including location, household income, and marital status. In addition, statistical tests were conducted to determine the associations between selected demographic characteristics and PTSD symptoms.

### PTSD Patterns by Location

There was a statistically significant association between respondents’ location and PTSD severity (Pearson’s chi-square test, p < 0.001). The distribution of PCL-5 scores differed substantially across Waretta, Gendabi, and Jorodom. Gendabi recorded the greatest proportion of respondents with very high PCL-5 scores. Of the 152 respondents from Gendabi, 64 (42.1%) scored above 60, while a further 32 (21.1%) scored between 45 and 59. Thus, approximately two-thirds of respondents from Gendabi (63.2%) had PCL-5 scores of 45 or higher. In contrast, Jorodom had no respondents with scores above 60, and the largest proportion of respondents in this location had relatively lower scores: 29 (33.0%) scored 15– 29, while 25 (28.4%) scored 0–14. Waretta showed an intermediate pattern. Among the 120 respondents, 14 (11.7%) had scores above 60, 30 (25.0%) scored 45–59**, and** 38 (31.7%) scored 30–44. Overall, the distribution indicates substantially greater PTSD symptom severity among respondents from Gendabi compared with those from Jorodom and Waretta (Table 3).

**Table 3.**
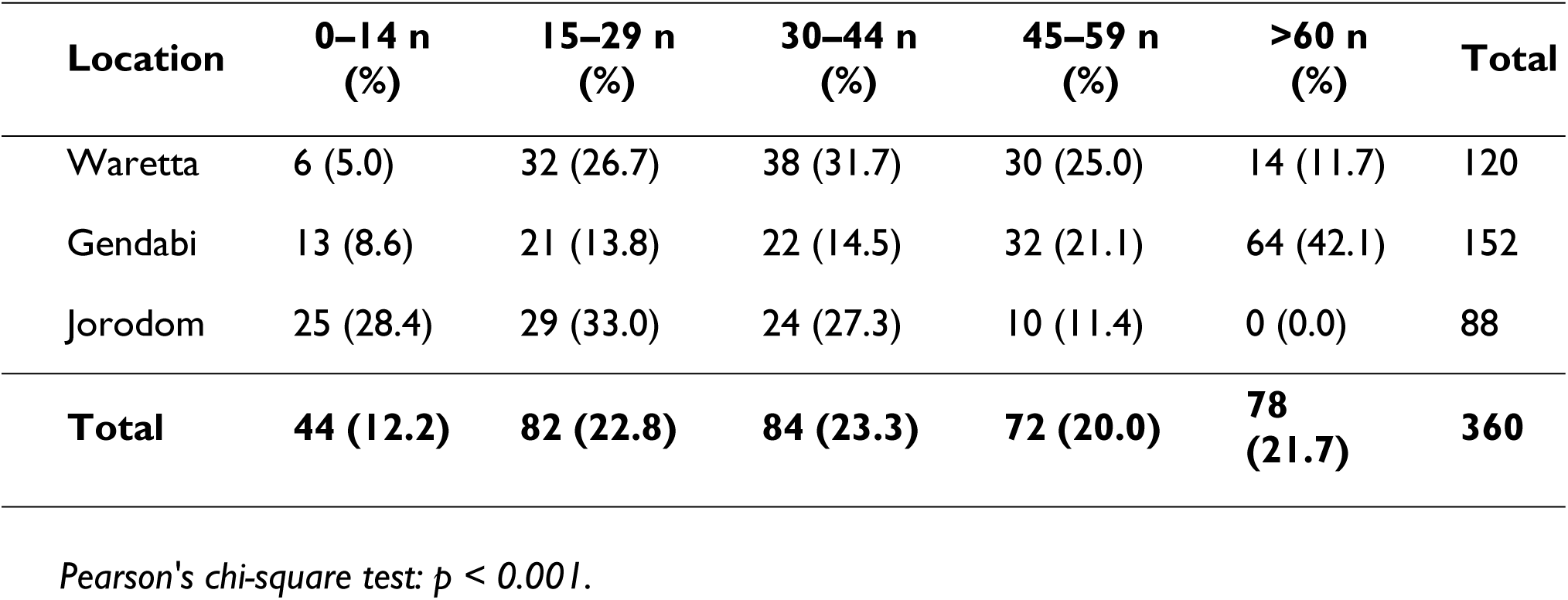
PTSD Severity by Location.

| Location | 0–14 n (%) | 15–29 n (%) | 30–44 n (%) | 45–59 n (%) | >60 n (%) | Total |
| --- | --- | --- | --- | --- | --- | --- |
| Waretta | 6 (5.0) | 32 (26.7) | 38 (31.7) | 30 (25.0) | 14 (11.7) | 120 |
| Gendabi | 13 (8.6) | 21 (13.8) | 22 (14.5) | 32 (21.1) | 64 (42.1) | 152 |
| Jorodom | 25 (28.4) | 29 (33.0) | 24 (27.3) | 10 (11.4) | 0 (0.0) | 88 |
| **Total** | **44 (12.2)** | **82 (22.8)** | **84 (23.3)** | **72 (20.0)** | **78 (21.7)** | **360** |

*Pearson's chi-square test: $p < 0.001$ .*

The marked geographical differences may reflect variation in the intensity of flood exposure, extent of property and livelihood losses, availability of social support, accessibility of health services, and effectiveness of local disaster response mechanisms. The particularly high proportion of severe scores in Gendabi suggests that location-specific contextual factors may have contributed to psychological vulnerability among flood survivors.

### PTSD Patterns by Household Income Level

Household income was significantly associated with PTSD severity (Pearson’s chi-square test, p = 0.011). Respondents from lower-income households generally had a greater concentration of severe PTSD scores than those from higher-income households. Among respondents earning less than TZS 50,000 per month, 34 (20.0%) had PCL-5 scores above 60, while 41 (24.1%) scored between 45 and 59. Similarly, among respondents earning TZS 50,000–150,000, 34 (26.0%) had scores above 60 and 24 (18.3%) scored between 45 and 59. In contrast, only 8 (16.7%) respondents in the TZS 150,000–300,000 income category and 2 (18.2%) of those earning more than TZS 300,000 had scores above 60. The highest-income group also had no respondents with scores between 45 and 59, although the relatively small sample size in this category (n = 11) should be considered when interpreting the findings. The findings indicate that economic circumstances were associated with PTSD severity. Lower-income households may have fewer financial resources to replace damaged property, restore livelihoods, access healthcare, and meet household needs following a disaster (Table 4).

**Table 4.** PTSD Severity by Household Income Level.

| Household income (TZS/month) | 0–14 n (%) | 15–29 n (%) | 30–44 n (%) | 45–59 n (%) | >60 n (%) | Total |
| --- | --- | --- | --- | --- | --- | --- |
| <50,000 | 11 (6.5) | 38 (22.4) | 46 (27.1) | 41 (24.1) | 34 (20.0) | 170 |
| 50,000–150,000 | 18 (13.7) | 27 (20.6) | 28 (21.4) | 24 (18.3) | 34 (26.0) | 131 |
| 150,000–300,000 | 11 (22.9) | 15 (31.3) | 7 (14.6) | 7 (14.6) | 8 (16.7) | 48 |
| >300,000 | 4 (36.4) | 2 (18.2) | 3 (27.3) | 0 (0.0) | 2 (18.2) | 11 |
| **Total** | **44 (12.2)** | **82 (22.8)** | **84 (23.3)** | **72 (20.0)** | **78 (21.7)** | **360** |

Pearson's chi-square test: $p = 0.011$ .

### PTSD Patterns by Marital Status

Marital status was significantly associated with PTSD severity (Pearson’s chi-square test, p = 0.002). The distribution of PCL-5 scores varied considerably across single, married, divorced/separated, and widowed respondents. Among the 235 married respondents, 64 (27.2%) had PCL-5 scores above 60, representing the largest number of severe cases across the marital-status categories. A further 43 (18.3%) scored between 45 and 59. Thus, approximately 45.5% of married respondents had scores of 45 or higher. Among widowed respondents, 6 (20.0%) had scores above 60 and 10 (33.3%) scored between 45 and 59. Therefore, more than half (53.3**%**) of widowed respondents had scores of 45 or higher. Divorced/separated respondents had relatively fewer severe cases in absolute numbers, with 3 (11.5%) scoring above 60 and 3 (11.5%) scoring between 45 and 59. Among single respondents, 5 (7.2%) had scores above 60 (Table 5).

**Table 5.** PTSD Severity by Marital Status.

| Marital status | 0–14 n (%) | 15–29 n (%) | 30–44 n (%) | 45–59 n (%) | >60 n (%) | Total |
| --- | --- | --- | --- | --- | --- | --- |
| Single | 10 (14.5) | 15 (21.7) | 23 (33.3) | 16 (23.2) | 5 (7.2) | 69 |
| Married | 28 (11.9) | 54 (23.0) | 46 (19.6) | 43 (18.3) | 64 (27.2) | 235 |
| Divorced/Separated | 4 (15.4) | 11 (42.3) | 5 (19.2) | 3 (11.5) | 3 (11.5) | 26 |
| Widowed | 2 (6.7) | 2 (6.7) | 10 (33.3) | 10 (33.3) | 6 (20.0) | 30 |
| **Total** | **44 (12.2)** | **82 (22.8)** | **84 (23.3)** | **72 (20.0)** | **78 (21.7)** | **360** |

*Pearson's chi-square test: $p = 0.002$ .*

### Factors Associated with PTSD Symptoms

**Table 6** presents the statistical analysis of demographic and socioeconomic factors associated with PTSD symptoms among flood victims in Hanang District. The analysis included gender, marital status, occupation, age, educational level, household income, and number of dependents. Statistical significance was assessed at the 5% significance level (p < 0.05). The findings show that gender was not significantly associated with PTSD symptoms among the respondents (χ² test, p = 0.314). This indicates that the level of PTSD symptoms did not differ significantly between male and female flood victims. In contrast, marital status was significantly associated with PTSD symptoms (χ² test, p = 0.002). This finding suggests that PTSD symptom severity differed according to marital status, with the descriptive results indicating a relatively greater burden of severe symptoms among married and widowed respondents compared with some other marital-status groups.

**Table 6.** Factors Associated with PTSD Symptoms among Flood Victims in Hanang District.

| Variable | Statistical measure | p-value |
| --- | --- | --- |
| Gender | Chi-square | 0.314 |
| Marital status | Chi-square | 0.002 |
| Occupation | Chi-square | 0.052 |
| Age | Regression coefficient = 0.019 | 0.001 |
| Educational level | Regression coefficient = 0.047 | 0.726 |
| Household income | Regression coefficient = -0.297 | 0.001 |
| Number of dependents | Correlation coefficient = -0.251 | 0.022 |

*Statistical significance was assessed at $\alpha = 0.05$ .*

Occupation was not statistically significantly associated with PTSD symptoms at the 5% level (χ² test, p = 0.052). Although the p-value was close to the conventional significance threshold, the result indicates insufficient statistical evidence to conclude that PTSD symptoms differed significantly according to occupation. Age demonstrated a positive and statistically significant association with PTSD symptoms (regression coefficient = 0.019, p = 0.001). This suggests that increasing age was associated with increasing PTSD symptom severity. Thus, older respondents may have experienced a greater psychological burden following the flooding. Similarly, educational level was not significantly associated with PTSD symptoms (regression coefficient = 0.047, p = 0.726). The result indicates that differences in respondents’ educational attainment did not explain significant differences in PTSD symptom severity within the study population (Table 6).

Household income showed a negative and statistically significant association with PTSD symptoms (regression coefficient = −0.297, p = 0.001). This indicates that respondents with higher household income tended to report lower levels of PTSD symptoms. The finding suggests that greater economic resources may have provided better capacity to cope with the consequences of flooding, including loss of property, disruption of livelihoods, and access to healthcare and other forms of support. Finally, the number of dependents was significantly and negatively correlated with PTSD symptoms (r = −0.251, p = 0.022). This indicates that respondents with a greater number of dependents tended to report lower PTSD symptom severity. Although this finding is contrary to the expectation that greater family responsibilities may increase psychological stress, it may reflect the protective role of family and social connectedness in the Hanang context. However, this finding should be interpreted cautiously because the analysis demonstrates an association rather than a causal protective effect (Table 6).

### Cognitive Factors Associated with PTSD Symptoms Among Flood Victims in Hanang District

**Table 7** presents the distribution of traumatic experiences reported by the 360 flood victims in Hanang District. The findings indicate that respondents were exposed to a wide range of traumatic events, either directly, as witnesses, or through learning about the events. Overall, the results demonstrate that direct exposure to floods and indirect exposure to other traumatic events were common, suggesting that PTSD risk in the study population may have been shaped by both personal experiences and exposure to trauma affecting other people in the community.

**Table 7:**
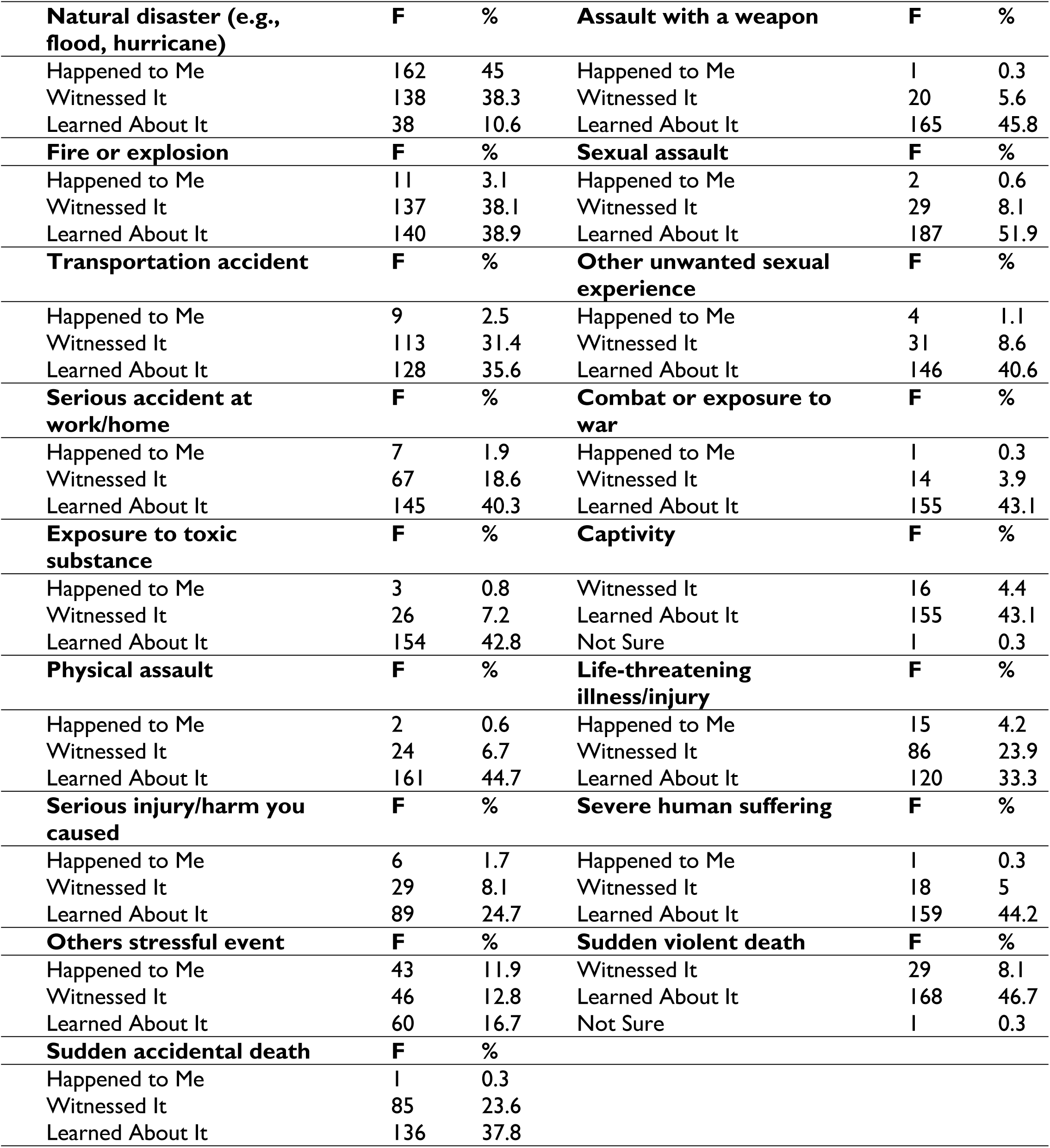
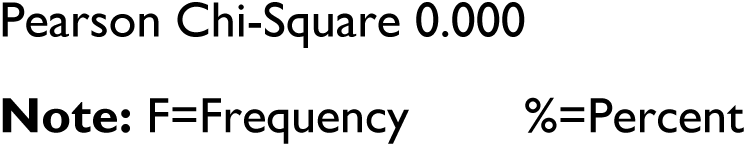
Cognitive factors contributing to the development of PTSD Symptoms Among Flood Victims in Hanang District.

The most frequently reported traumatic experience was natural disaster, particularly flooding. A total of 162 respondents (45.0%) reported that the disaster happened directly to them, while 138 (38.3%) witnessed it and 38 (10.6%) learned about it. This indicates that flooding was the predominant traumatic experience among respondents and represents an important potential contributor to PTSD symptoms in the study population. The high proportion of respondents who directly experienced or witnessed the flood reflects the widespread nature of the Hanang disaster and suggests that psychological consequences may extend beyond individuals who directly suffered physical or material losses. Other traumatic experiences were predominantly encountered indirectly through witnessing or learning about them. For example, only 1 respondent (0.3%) reported experiencing assault with a weapon directly, compared with 20 (5.6%) who witnessed such an event and 165 (45.8%) who learned about it. Similarly, for fire or explosion, 11 respondents (3.1%) experienced the event directly, while 137 (38.1%) witnessed it and 140 (38.9%) learned about it. These findings demonstrate that although direct exposure to some forms of trauma was relatively uncommon, information about traumatic events was widespread within the community (Table 7).

A similar pattern was observed for sexual assault and other unwanted sexual experiences. Only 2 respondents (0.6%) reported direct sexual assault, whereas 29 (8.1%) witnessed it and 187 (51.9%) learned about it. For other unwanted sexual experiences, 4 respondents (1.1%) reported direct experience, 31 (8.6%) witnessed such experiences, and 146 (40.6%) learned about them. Although these events were less commonly experienced directly, the high proportion of respondents who had learned about them indicates substantial community-level exposure to distressing information. For transportation accidents, 9 respondents (2.5%) experienced an accident directly, 113 (31.4%) witnessed an accident, and 128 (35.6%) learned about such an event. Likewise, serious accidents occurring at home or at work were directly experienced by 7 respondents (1.9%), witnessed by 67 (18.6%), and learned about by 145 (40.3%). These findings suggest that accidental and potentially life-threatening events formed an important component of the broader trauma exposure experienced by the respondents (Table 7).

Exposure to combat or war was relatively uncommon as a direct experience, with only 1 respondent (0.3%) reporting direct exposure. However, 14 respondents (3.9%) had witnessed such events and 155 (43.1%) had learned about them. Similarly, exposure to toxic substances was reported directly by only 3 respondents (0.8%), while 16 (4.4%) witnessed such exposure and 155 (43.1%) learned about it. This pattern again highlights the importance of indirect exposure to traumatic events within the community. For captivity, 26 respondents (7.2%) reported witnessing such experiences and 154 (42.8%) had learned about them, while only 1 respondent (0.3%) was uncertain about the experience. Physical assault was directly experienced by 2 respondents (0.6%), witnessed by 24 (6.7%), and learned about by 161 (44.7%). These findings indicate that information about traumatic events was substantially more common than direct personal exposure (Table 7).

Similarly, life-threatening illness or injury was directly experienced by 15 respondents (4.2%), witnessed by 86 (23.9%), and learned about by 120 (33.3%). Serious injury or harm caused to another person was reported directly by 6 respondents (1.7%), witnessed by 29 (8.1%), and learned about by 89 (24.7%). Severe human suffering was directly experienced by 1 respondent (0.3%), witnessed by 18 (5.0%), and learned about by 159 (44.2%). These results demonstrate that respondents were exposed not only to the flood itself but also to substantial information about suffering and other traumatic experiences within their communities. The findings for other stressful events were also notable. Forty-three respondents (11.9%) reported experiencing such events directly, 29 (8.1%) witnessed them, and 168 (46.7%) learned about them. For sudden violent death, 46 respondents (12.8%) witnessed such an event, while 60 (16.7%) learned about it. Only 1 respondent (0.3%) reported direct experience. In relation to sudden accidental death, 1 respondent (0.3%) experienced it directly, 85 (23.6%) witnessed it, and 136 (37.8%) learned about it (Table 7).

Overall, the results show a consistent pattern in which direct exposure was highest for natural disasters/flooding, whereas indirect exposure through witnessing or learning about traumatic events was more common for most other types of trauma. This distinction is important because psychological responses to disasters may not be limited to those directly affected. Witnessing traumatic events or repeatedly hearing about severe injury, death, violence, or suffering within a close-knit community may also contribute to psychological distress. The reported Pearson chi-square value of 0.000 indicates a statistically significant association between the assessed traumatic/cognitive exposure factors and PTSD manifestations at the conventional 5% significance level. Therefore, the null hypothesis of no association is rejected. The findings suggest that the pattern and type of exposure to traumatic experiences were significantly related to PTSD outcomes among flood victims in Hanang District (Table 7).

## Discussion

This study found a substantial burden of post-traumatic stress symptoms among survivors of the December 2023 Hanang landslide-flood disaster approximately one year after the event. The mean PCL-5 score was 39.2 (SD = 20.6), and approximately 45% of participants had scores ≥45, indicating a high level of PTSD symptomatology in this population. The persistence of substantial symptoms one year after the disaster is consistent with evidence that psychological consequences of disasters can extend well beyond the acute emergency period, particularly when survivors experience severe trauma, displacement, economic losses, disruption of social networks, and continuing stressors (Goldmann & Galea, 2014; Neria et al., 2008; North & Pfefferbaum, 2013). The PCL-5 is a validated self-report instrument for assessing PTSD symptoms corresponding to DSM-5 criteria, but a screening score does not by itself establish a clinical diagnosis. A recent study found considerable variation in PTSD prevalence across populations and traumatic events, emphasizing that estimates are influenced by the type and severity of trauma, characteristics of the affected population, and assessment method (Schincariol et al., 2024). Thus, the relatively high level of symptoms observed in Hanang may reflect the severity of the landslide-flood disaster, the multiplicity of traumatic experiences, continuing socioeconomic disruption, and the rural context in which recovery occurred.

### Geographic variation in PTSD manifestations

A significant geographical difference in PTSD severity was observed across Gendabi, Jorodom, and Waretta (p < 0.001), with Gendabi reporting the greatest proportion of severe symptoms. This finding suggests that psychological consequences may not be distributed uniformly within a disaster-affected district. Differences between communities may reflect variation in the intensity of exposure, deaths and injuries, destruction of housing and livelihoods, displacement, availability of assistance, social support, and perceived safety during recovery. The finding is consistent with disaster research showing that the severity of exposure and circumstances surrounding and following the traumatic event are important determinants of psychological outcomes. A meta-analysis by Brewin et al. (2000) found that trauma severity, lack of social support, and additional life stress were among the factors with relatively stronger associations with PTSD compared with several pre-trauma characteristics. Similarly, evidence from disaster research indicates that psychological consequences are shaped by both characteristics of the disaster and the social environment in which recovery occurs (Goldmann & Galea, 2014). The geographical differences identified in Hanang may therefore reflect a combination of initial disaster exposure and post-disaster recovery conditions. Communities experiencing greater destruction may face more persistent reminders of the disaster, greater economic losses, disrupted social networks, and continuing uncertainty. These factors may maintain psychological distress even after the immediate physical danger has passed. From a public health perspective, the geographical variation has implications for the allocation of disaster mental health resources. Mental health interventions should not necessarily be distributed equally across all affected communities. Instead, areas with greater psychological burden should receive more intensive screening, psychosocial outreach, referral services, and follow-up.

### Socioeconomic vulnerability and PTSD

Household income was significantly associated with PTSD severity, and higher household income was independently associated with lower PCL-5 scores (β = −0.297, p = 0.001). This finding emphasizes the close relationship between economic recovery and psychological recovery following disasters. Disasters can produce substantial and prolonged economic consequences through destruction of homes, agricultural land, livestock, businesses, infrastructure, and other productive assets. In predominantly rural communities such as Hanang, where agriculture and livestock constitute important sources of livelihood, loss of productive assets may substantially reduce household capacity to recover. Continuing financial insecurity can subsequently become a source of chronic stress rather than allowing the traumatic experience to become a past event.

This interpretation is consistent with evidence showing that socioeconomic circumstances, additional life stress, and inadequate social resources contribute to post-traumatic psychological outcomes (Brewin et al., 2000; Goldmann & Galea, 2014). The World Health Organization also recognizes poverty, resource insecurity, loss, and disruption of essential services as factors that can increase mental health risks during emergencies. Several mechanisms may explain the observed association. First, households with limited resources may have difficulty reconstructing homes and replacing lost assets. Second, economic constraints may reduce access to healthcare and psychological services. Third, prolonged unemployment, food insecurity, and inadequate housing may maintain chronic stress. Finally, continuing economic losses may reinforce perceptions of helplessness and lack of control. The protective association between income and PTSD symptoms should therefore not be interpreted as evidence that income itself prevents PTSD. Rather, greater economic resources may provide survivors with more opportunities to restore housing, livelihoods, social functioning, and a sense of control. Conversely, persistent poverty may prolong exposure to post-disaster stressors. These findings support an important principle for disaster recovery in Tanzania: mental health recovery should be integrated with livelihood recovery. Psychosocial interventions implemented without addressing severe economic hardship may be insufficient because material circumstances may continue to generate stress and reinforce perceptions of insecurity.

### Age and PTSD severity

Older age was positively associated with PTSD severity (β = 0.019, p = 0.001). Although the regression coefficient was relatively small, the association suggests that older survivors may require particular attention during post-disaster mental health screening. Older adults may experience disaster-related losses differently from younger adults. They may have greater difficulty recovering from physical injury, replacing lost productive assets, relocating, or adapting to changes in their living environment. They may also have fewer economic opportunities for rebuilding livelihoods. These factors can interact with disaster exposure and social support to influence psychological recovery. The relationship between age and PTSD is not necessarily consistent across all disasters. Brewin et al. (2000) found that age and other demographic characteristics showed variable associations across trauma-exposed populations, whereas characteristics such as trauma severity, social support, and additional life stress demonstrated more consistent effects. Therefore, age should be regarded as one component of a broader vulnerability profile rather than as an independent explanation for PTSD. The finding nonetheless suggests that post-disaster programmes should ensure that older adults are not overlooked during mental health screening. Particular attention may be required for older people who simultaneously experience bereavement, physical limitations, loss of property, reduced income, and disruption of established social networks.

### Marital status and PTSD

Marital status was significantly associated with PTSD manifestations (p = 0.002). Married participants represented a substantial proportion of those with severe symptoms, while widowed and divorced participants also demonstrated considerable symptom burden. The relationship between marital status and PTSD is potentially complex. Marriage may provide emotional, social, and practical support following a disaster, but it can also involve substantial responsibilities for dependants and household recovery. For married survivors, the psychological consequences of the disaster may therefore include concern about the safety and welfare of spouses, children, and other family members. For widowed and divorced survivors, the absence of a partner may reduce access to immediate emotional and practical support. In rural communities, where households and extended family networks may constitute important sources of social protection, disruption of these relationships may be particularly consequential. The broader disaster literature supports the importance of social support and post-disaster stress in shaping PTSD outcomes (Brewin et al., 2000; Goldmann & Galea, 2014). Consequently, marital status should not be interpreted simply as a biological or demographic risk factor; rather, it may operate through social support, household responsibilities, bereavement, and access to practical resources.

### Trauma exposure and PTSD

One of the strongest findings of this study was the significant association between trauma exposure and PTSD symptoms (p < 0.001). Direct experience of the flood was reported by 45% of participants, while 38.3% witnessed the disaster and 10.6% learned about the disaster affecting others. Participants also reported exposure to accidents, serious injuries, deaths, violence, and severe human suffering. These findings are consistent with the DSM-5-TR conceptualization of traumatic exposure, which recognizes direct experience, witnessing traumatic events, learning about traumatic experiences affecting close others under specified circumstances, and repeated exposure to traumatic details as relevant forms of exposure (American Psychiatric Association, 2022). The findings also support evidence that severity and multiplicity of trauma exposure are important determinants of post-traumatic symptoms. A meta-analysis of PTSD risk factors found that trauma severity and additional life stress were among the more important predictors of PTSD across trauma-exposed adults (Brewin et al., 2000). Similarly, research following disasters indicates that the psychological burden tends to increase with greater exposure to death, injury, loss, displacement, and other severe consequences of the event (Goldmann & Galea, 2014; Tang et al., 2017). The findings from Hanang therefore suggest that disaster mental health screening should extend beyond individuals who sustained physical injuries or whose homes were destroyed. Survivors who witnessed deaths, serious injuries, rescue operations, or severe suffering may also experience substantial psychological consequences.

### Cognitive processing and PTSD

The association between trauma exposure and PTSD symptoms is particularly relevant when interpreted through cognitive models of PTSD. Ehlers and Clark (2000) proposed that persistent PTSD develops when traumatic experiences are processed in a way that produces a persistent sense of current threat. This may occur through excessively negative appraisals of the trauma or its consequences and through trauma memories that are poorly contextualized and easily triggered by reminders. The Hanang findings are compatible with this framework. Although the quantitative assessment primarily measured trauma exposure rather than validated cognitive constructs, participants were exposed to circumstances likely to generate persistent perceptions of threat, helplessness, loss, and uncertainty. Continued exposure to reminders of destroyed homes, damaged environments, lost livelihoods, bereavement, and unpredictable weather may maintain the perception that danger remains present. This distinction is important because trauma exposure and cognitive processing should not be treated as synonymous concepts. Directly experiencing a flood, witnessing a death, or learning about a traumatic event represents exposure to trauma. Cognitive factors refer to how individuals interpret, remember, and assign meaning to those experiences. The present study provides evidence for the importance of trauma exposure but does not establish that specific maladaptive cognitions caused PTSD. Future studies should therefore directly measure cognitive constructs such as negative beliefs about the self and world, perceived threat, guilt, shame, helplessness, and trauma-related appraisals. Such measures would allow a more rigorous test of the cognitive mechanisms proposed by Ehlers and Clark (2000).

### Direct, witnessed, and indirect trauma exposure

The substantial proportion of participants who witnessed the disaster or learned about traumatic experiences highlights the importance of considering community-level trauma. In closely connected communities, the consequences of a disaster are not restricted to individuals who were physically injured. Deaths, injuries, displacement, destruction of homes, and loss of livelihoods may affect entire families and communities. Repeated conversations about deaths and losses, exposure to grieving relatives, and continued observation of damaged environments may create persistent reminders of the traumatic event. These experiences may be particularly important when they occur alongside direct exposure. However, the present findings should not be interpreted as demonstrating that all forms of indirect exposure independently cause PTSD. The DSM-5-TR provides specific criteria for qualifying indirect exposure, and learning about trauma affecting unrelated community members is not equivalent diagnostically to learning about trauma affecting a close family member or friend (American Psychiatric Association, 2022). This distinction should be retained in the manuscript to avoid overextending the diagnostic definition of trauma exposure.

### Interaction between socioeconomic stress and trauma processing

The simultaneous associations of PTSD with trauma exposure and household income suggest that psychological recovery may be influenced by both the initial traumatic event and the conditions that follow it. Economic hardship may maintain distress by making the consequences of the disaster continuously visible in survivors’ daily lives. For example, the loss of farmland, livestock, housing, or income may reinforce perceptions of helplessness, insecurity, or an uncertain future. In contrast, successful livelihood restoration may help survivors regain a sense of control and predictability. This interpretation is consistent with cognitive models that emphasize the role of ongoing threat and negative appraisals in maintaining PTSD (Ehlers & Clark, 2000), as well as disaster research showing that additional life stress and inadequate social resources can contribute to persistent psychological symptoms (Brewin et al., 2000; Goldmann & Galea, 2014). The implication is that psychological interventions should not be implemented independently of broader recovery programmes. Trauma-focused treatment may be more effective when accompanied by interventions addressing housing, livelihoods, social protection, safety, and access to essential services.

### Implications for mental health services in Tanzania

The findings have important implications for disaster preparedness and mental health service delivery in Tanzania. Disaster responses frequently prioritize rescue, shelter, food, water, sanitation, injuries, and infectious disease prevention. These remain essential, but the present findings demonstrate the need for mental health interventions to continue into the recovery phase. The WHO emphasizes that emergencies, including natural disasters, can disrupt health services and increase exposure to loss, poverty, resource insecurity, and other stressors that affect mental health. Mental health therefore needs to be integrated across the disaster management continuum, from preparedness and emergency response through recovery and reconstruction. At the primary healthcare level, health workers could be trained to identify PTSD symptoms, provide psychoeducation and basic psychosocial support, and establish referral pathways for individuals requiring specialized care. The PCL-5 can support systematic assessment of PTSD symptoms, although positive screening results should be followed by appropriate clinical assessment where possible (Blevins et al., 2015). Community-based approaches are particularly relevant in rural settings. Community health workers, local leaders, social service providers, and other trusted community structures may help identify individuals experiencing persistent distress and facilitate referral. Such interventions should remain trauma-informed and culturally appropriate and should distinguish normal responses to bereavement and disaster-related stress from mental disorders requiring clinical treatment.

The findings support consideration of trauma-focused psychological interventions, particularly for survivors with persistent or severe symptoms. Cognitive approaches are relevant because the theoretical model underpinning this study proposes that persistent PTSD is partly maintained by negative appraisals and a continuing sense of threat (Ehlers & Clark, 2000). Interventions should therefore address trauma-related beliefs about safety, helplessness, guilt, responsibility, loss, and the future where these beliefs are clinically relevant. At the same time, interventions should not focus exclusively on cognition when survivors continue to experience severe material insecurity. Psychological treatment should be complemented by interventions that address housing, livelihoods, social support, and other continuing stressors.

The significant association between household income and PTSD severity supports greater integration of mental health, livelihood restoration, and social protection within disaster recovery policies. Rebuilding homes and infrastructure alone may not be sufficient if survivors remain unable to restore livelihoods or meet basic household needs. The geographical variation in PTSD also supports differentiated resource allocation. Mental health resources should be directed according to both physical disaster impact and observed psychological need. Communities with high levels of severe PTSD symptoms may require more intensive screening, outreach, referral, and follow-up. The persistence of symptoms approximately one year after the disaster further indicates that disaster mental health services should extend beyond the immediate emergency period. Recovery programmes should include mechanisms for continued psychological monitoring and referral.

### Strengths and limitations

A major strength of this study is its community-based assessment approximately one year after a severe landslide-flood event. This timing provides insight into psychological consequences beyond the acute emergency phase. The use of the PCL-5 provides a standardized approach to measuring PTSD symptoms, and the study also considered demographic, socioeconomic, geographical, and trauma-exposure factors. The mixed-methods approach provides an additional strength by allowing quantitative patterns to be considered alongside survivors’ accounts of their experiences. This can provide a richer understanding of how disaster exposure and post-disaster circumstances influence psychological well-being.

Several limitations should be considered. First, the cross-sectional design prevents causal inference. It cannot be determined whether the identified cognitive or socioeconomic factors preceded PTSD symptoms or were themselves consequences of the disaster and subsequent psychological distress. Longitudinal studies beginning shortly after the disaster would provide stronger evidence regarding PTSD trajectories. Second, trauma exposure and PTSD symptoms were self-reported and may therefore be affected by recall or reporting bias, particularly because interviews occurred approximately one year after the disaster. Third, the PCL-5 is a screening instrument rather than a diagnostic interview; therefore, the study should avoid describing participants with scores ≥45 as having clinically confirmed PTSD. Fourth, the study was conducted in three wards of Hanang District, which may limit generalizability to other regions or disaster contexts in Tanzania. Finally, although the manuscript describes “cognitive factors,” the quantitative instrument appears to assess trauma exposure more strongly than specific cognitive processes. Future research should include validated measures of trauma-related cognitions, perceived threat, negative beliefs, guilt, and maladaptive appraisals. This would allow more direct testing of the cognitive model of PTSD proposed by Ehlers and Clark (2000).

## Conclusion

PTSD represents a substantial mental health burden among flood victims in Hanang District, with a considerable proportion of survivors continuing to experience moderate to severe PTSD symptoms one year after the disaster. The findings demonstrate that PTSD manifestations varied significantly by geographical location, household income, marital status, and age, while gender and educational level were not significantly associated with PTSD symptoms. Lower household income and increasing age were associated with greater PTSD severity, whereas the number of dependents showed an inverse association with PTSD symptoms. In addition, exposure to traumatic events—including direct experience of the flood, witnessing traumatic events, and learning about traumatic experiences affecting others—was significantly associated with PTSD manifestations. These findings highlight that the psychological consequences of disasters extend beyond individuals directly affected and may affect the wider community through indirect exposure to trauma. Effective disaster recovery in Hanang therefore requires the integration of mental health services into humanitarian and disaster-response programmes, including routine PTSD screening, trauma-informed care, community-based cognitive behavioural therapy, psychosocial support, and livelihood recovery interventions. Strengthening community resilience, social support systems, and mental health preparedness within national disaster risk-reduction strategies will also be essential for reducing the long-term psychological consequences of future disasters.

## Data Availability

The minimal data set and accompanying code, where generated, is available at the attached filles as Supporting Documents

